# Genome-Wide Significance Reconsidered: Low-Frequency Variants and Regulatory Networks in Autism

**DOI:** 10.64898/2026.02.11.26346090

**Authors:** Marla Mendes, Worrawat Engchuan, Sydney Thompson, Xiaopu Zhou, Nickie Safarian, Desmond Zeya Chen, Brett Trost, Nelson Bautista Salazar, Clement Ma, Bhooma Thiruvahindrapuram, Jacob Vorstman, Stephen W. Scherer, Elemi Breetvelt

**Affiliations:** The Centre for Applied Genomics, The Hospital for Sick Children, Toronto, ON, Canada; Genetics and Genome Biology Program, The Hospital for Sick Children, Toronto, ON, Canada; Molecular Medicine Program, The Hospital for Sick Children, Toronto, ON, Canada; Division of Biostatistics, Dalla Lana School of Public Health, University of Toronto, Toronto, ON, Canada; Centre for Addiction and Mental Health, Toronto, Ontario, Canada; Department of Molecular Genetics, University of Toronto, Toronto, ON, Canada; McLaughlin Centre, University of Toronto, Toronto, ON, Canada; Department of Psychiatry, Leiden University Medical Center, Leiden, the Netherlands

**Keywords:** Significant Threshold, Autism, Minor Allele Frequency, GWAS, Zinc Finger genes, Low Frequency Variants (LFVs)

## Abstract

Low-frequency variants (LFVs), defined by minor allele frequencies (MAF) of 1–5%, occupy the gap between common and rare variants in both frequency and effect size. The conventional genome-wide association study (GWAS) significance threshold (5×10⁻⁸) is overly conservative for LFVs, which account for more than 25% of variants in GWAS. This limitation may obscure meaningful associations in highly heritable yet genetically complex disorders such as autism spectrum disorder (ASD). We hypothesize that the scarcity of significant LFVs in ASD GWAS reflects statistical constraints rather than a true lack of association. To address this, we derived a MAF-specific genome-wide significance threshold using linkage disequilibrium–informed simulations applied to ASD GWAS summary statistics, identifying 2.03×10⁻⁷ as optimal. Applying this threshold revealed three novel LFVs mapping to zinc finger proteins (ZNF420, ZNF781) and known ASD-related genes (KMT2E, PRKDC, MCM4). Enrichment analyses suggested their function in nervous system development and gene regulation. Our findings highlight the contribution of LFVs to ASD risk and underscore the importance of frequency-aware association strategies.

## Introduction

Genetic studies have greatly advanced our understanding of the genomic architecture underlying neurodevelopmental and psychiatric conditions ^1–6^. Approaches spanning the full spectrum of allele frequencies have been instrumental in these advances ^7–10^. At one end, large-scale genome-wide association studies (GWAS) have identified numerous common variants (minor allele frequency [MAF] >5%) associated with modest effects on risk^11^. At the other, analyses of rare variants (MAF <1%), including studies of copy number variants (CNVs) and family-based sequencing, have revealed genes with large effect sizes and critical roles in neurodevelopmental processes ^12,13^.

Despite this progress, a gap remains for variants in the intermediate frequency range. Low-frequency variants (LFVs; MAF 1–5%) account for over a quarter of all single nucleotide polymorphisms (SNPs) represented in GWAS, yet few have been significantly associated with neurodevelopmental or psychiatric phenotypes^11^. This gap limits our understanding of the genetic architecture and biological mechanisms contributing to these disorders^14^.

Autism spectrum disorder (ASD) exemplifies this challenge. ASD is a group of highly heritable neurodevelopmental conditions for which genetic risk arises from variants across the entire allele frequency spectrum^7,10,15,16^. Common variant GWAS have identified several associated loci, but these account for only a modest proportion of heritability ^11^. Conversely, rare variant analyses have uncovered high-impact mutations in a subset of genes, yet together they explain only a fraction of genetic liability ^10^. The absence of robust associations for low-frequency variants highlights an underexplored component of ASD risk and a broader discovery gap shared with other psychiatric disorders, such as schizophrenia ^9^.

The analysis of low-frequency variants (LFVs) using GWAS, particularly in the context of binary traits such ASD is challenging. While association methods developed for common variants, such as logistic regression, can be applied to LFVs when sample sizes are sufficiently large, their performance is affected by calibration issues and reduced statistical power^17^. For example, the logistic regression Wald test is known to be overly conservative in case-control studies involving LFVs^18^. Overall, the calibration and power of logistic regression-based approaches for LFvar analysis in case-control settings remain insufficiently characterized.

We propose that the lack of genome-wide significant findings for LFVs likely stems from statistical constraints rather than a true absence of association. Simply put, given that robust and replicated association has been found for many variants in the common and in the rare frequency range, it is highly unlikely that association does not exist in the low frequency variant range. Since LFVs exhibit lower variation (limited amount of information that can be obtained from a SNP due to its MAF), they provide less Fisher information and yield higher standard errors for effect size estimates compared to common variants, requiring unfeasibly large sample sizes to provide sufficient statistical power when analyzed using the same conservative significance threshold^19^.

Greater attention to LFVs could help bridge the heritability gap observed in autism, as these variants may hold a significant and previously underappreciated role in shaping the genomic architecture of neurodevelopmental disorders. Moreover, our approach to deriving MAF-specific significance thresholds is generalizable and could enhance discovery in other complex traits studied through GWAS, where LFVs are similarly underpowered yet potentially impactful. In this study, we introduce MAF-specific significance thresholds tailored to LFVs, while applying Bonferroni correction to better balance the detection of non-effect variants (false positives) while reducing the likelihood of missing true-effect variants (false negatives). Using these more appropriate thresholds, we identify three novel loci associated with autism.

## Methods

### Summary Statistics

The summary statistics used in this study were obtained from the most recent Autism Spectrum Disorder (ASD) GWAS^11^, and are available for download at the Psychiatric Genomics Consortium website^20^. The frequency information can be obtained after an access request with ASD Data Access Committee (DAC). This data includes information from 9,060,460 SNPs. Of these, 2,336,259 (25.78%) have a Minor Allele Frequency (MAF) between 1% and 5% (based on 18,381 cases and 27,969 controls together), categorizing them as Low-Frequency variants (LFvar), while 6,720,996 have a MAF greater than 5% (74.18% of all GWAS SNPs).

### Significance threshold inference based on Bonferroni correction

To establish a more appropriate significance threshold for LFVs, we applied a Bonferroni correction using the number of *effectively independent* SNPs as the denominator, rather than the total number of variants tested. This accounts for the correlation structure among SNPs introduced by linkage disequilibrium (LD), which inflates the number of nominal tests if not adjusted for. Estimating the number of independent tests is a well-established strategy to maintain control of the family-wise error rate while improving statistical power, particularly in the context of whole-genome sequencing data where variant density is high (Figure 1).

**Figure 1.**
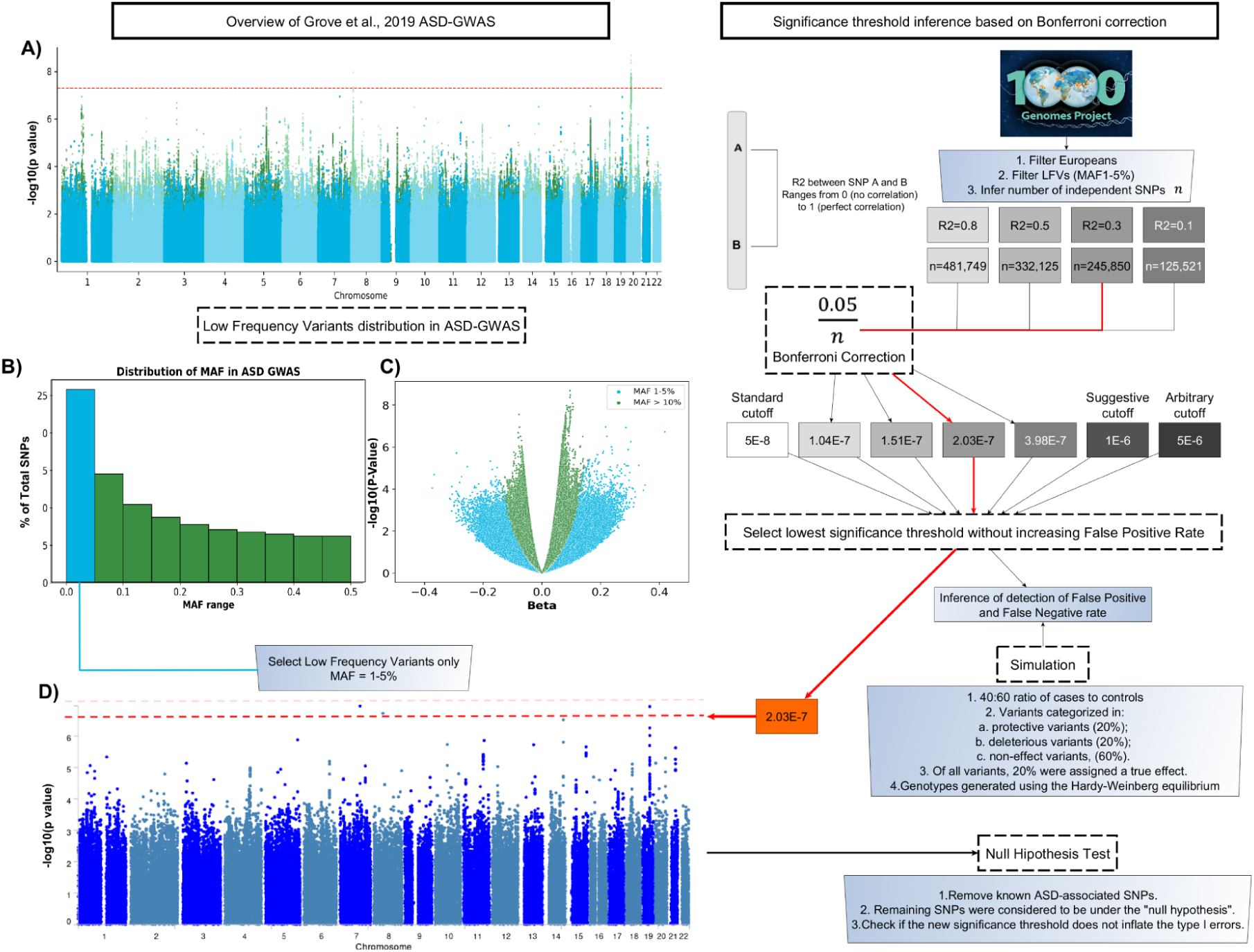
Workflow of the applied analysis. A) Genome-wide association study (GWAS) Manhattan plot adapted from Grove et al. (2019), showing the summary statistics used in the present analysis. LFVs are highlighted in blue, and variants with MAF > 5% are shown in green. The red dashed line indicates the conventional genome-wide significance threshold (P = 5 × 10⁻⁸). B) Distribution of ASD-GWAS variants across MAF categories, with the LFV range highlighted in light blue. C) Relationship between effect sizes (β) and association P-values for ASD-GWAS variants, highlighting LFVs in light blue. D) Manhattan plot of LFVs from the ASD-GWAS, with the red line indicating the adapted genome-wide significance threshold used in this study.

To empirically determine the number of independent SNPs, we conducted an LD pruning analysis across a range of LD thresholds (0.1 to 0.5) using the PLINK^21^ command --indep-pairwise 100 10^22,23^ <SELECTED R2>. Because LD patterns vary by ancestry, we used European samples from the 1000 Genomes Project^24^ as a reference population to ensure that the estimated LD structure was appropriate for the ASD discovery dataset. This analysis allowed us to identify a range of plausible correction factors for multiple testing, enabling a more tailored and statistically genome-wide significance threshold for low-frequency variants. This command applies a sliding window approach, evaluating 100-SNP windows and shifting by 10 SNPs at a time. Within each window, pairs of SNPs with LD (R²) greater than the selected threshold are identified, and one SNP from each high-LD pair is removed^21^. This process yields a pruned set of approximately independent variants, which we used as the denominator in the Bonferroni formula:

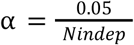

where *Nindep* is the number of LD-pruned variants. This adjustment provides a more realistic correction for multiple testing. Our final threshold selection was informed by simulation analyses, where we explicitly evaluated the detection of non-effect variants and the risk of missing true-effect variants under different LD cutoffs (False Positives and False Negatives, respectively. See Results and Figure 2).

**Figure 2.**
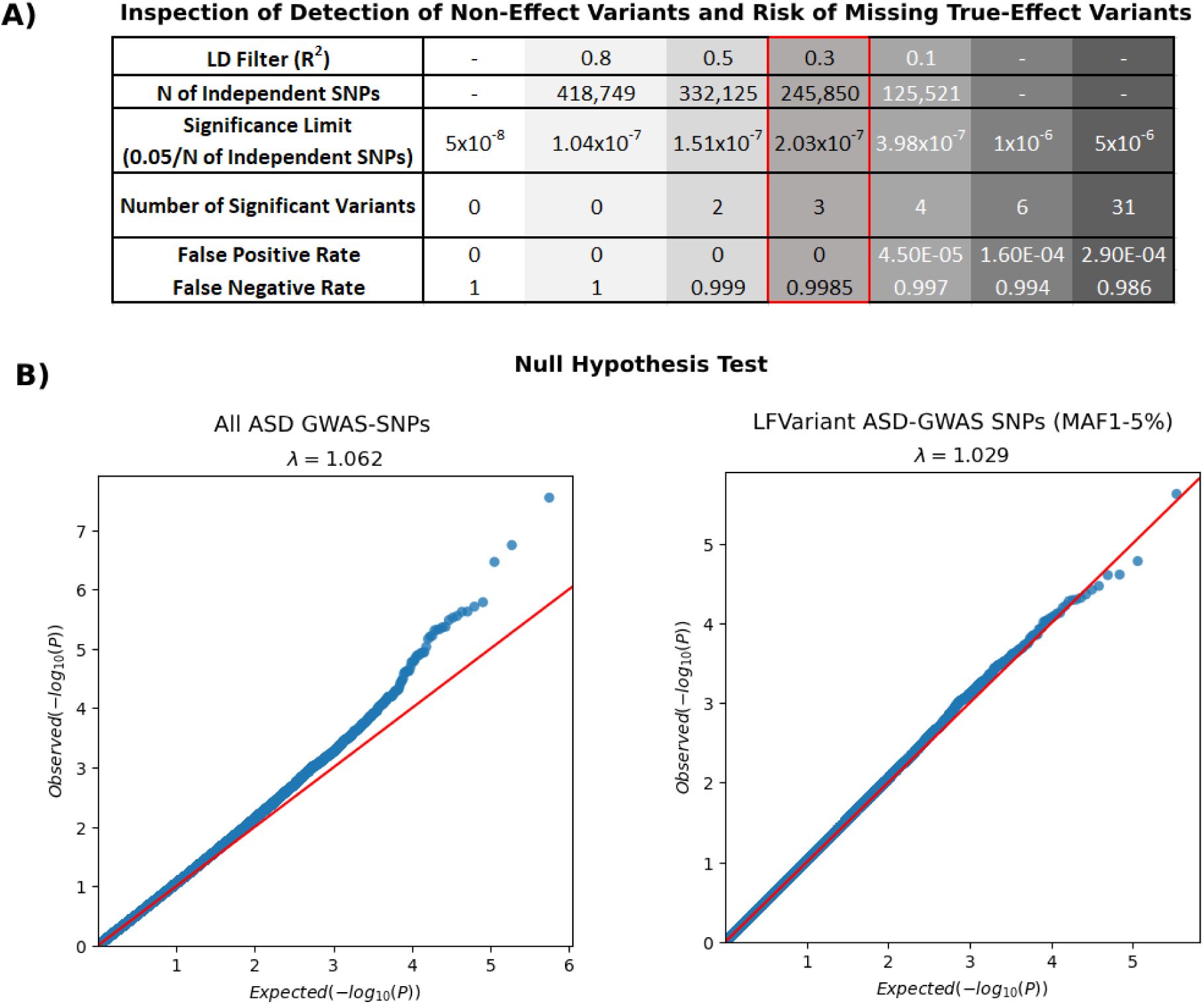
Appropriate Significance Threshold for LFVs. A) Summary of significance thresholds evaluated, including the number of significant SNPs and corresponding proportions of non-effect variants detected and true-effect variants missed. The red box highlights the final significance threshold selected in this study. B) Quantile-quantile (Q-Q) plots for all GWAS-SNPs compared with LFVs GWAS (MAF=1-5). Variants were filtered based on linkage disequilibrium (LD) at R² ≤ 0.3 and excluded if located within 1 megabase upstream or downstream of ASD-associated genes from the SFARI database^28^.

### Simulation

We simulated 1000 SNPs^25,26^ across 2000 individuals, with a case:control ratio of 40:60, reflecting common GWAS sampling structures. Genotypes were generated under a Hardy-Weinberg Equilibrium (HWE) assumption using allele frequencies randomly sampled from the range 0.01 to 0.05, consistent with LFVs. To simulate realistic differences in allele frequencies between cases and controls, we introduced random perturbations to the baseline of LFVs by applying shifts sampled from empirically derived normal distributions. These noise parameters, modeled with small positive means and variances, reflect natural variability in observed allele frequency estimates due to population-level factors. For protective variants in cases, and deleterious variants in controls, we reduced the MAFs by a random amount drawn from a mildly right-skewed normal distribution (mean = ∼4.6%, SD = ∼3%). Additional small-magnitude noise sampled from a separate distribution (mean = 0, SD = ∼0.5%) was added to both allele frequency arms to capture minor stochastic fluctuations.

SNPs were divided into three groups: 20% with protective effect (higher frequency in controls), 20% with deleterious effect (higher frequency in cases), and 60% with no effect. A additive disease model was assumed, and relative risk across genotypes was implicitly modeled by shifting allele frequencies between groups. Although HWE was assumed in generating the baseline population, we acknowledge that Hardy-Weinberg Disequilibrium (HWD) may occur in real-world datasets, warranting a careful check of HWE prior to any future analysis using our framework.

We defined the false positive rate as the proportion of non-effect SNPs with p-values below the significance threshold, and the false negative rate as the proportion of true-effect SNPs with p-values above the threshold. The script used to perform this simulation is available upon request.

### Assessment of False Positive Rate Under the Null Hypothesis

To assess whether our thus defined new significance threshold does not inflate the proportion of false positives, we performed a test under the null hypothesis: “none of the variants is associated with ASD”, using the LFVs on the published ASD-GWAS results ^11^. To this end, we first applied LD pruning with the selected threshold (R² = 0.3) to reduce correlation among variants. Next, we excluded all SNPs located within ±1 megabase of ASD-associated genes curated in the SFARI Gene database (all categories, total of 1,177 genes at 2024), ensuring the remaining variants were unlikely to be involved in ASD risk and could be assumed to follow the null hypothesis. While this approach aims to remove known ASD-associated loci, we acknowledge that our understanding of ASD genetics is still evolving and remains incomplete. Consequently, it is possible that some variants in the filtered set are truly associated with ASD but have not yet been identified. This makes our approach inherently conservative, such variants would be misclassified as false positives, potentially leading to an overestimation of the false positive rate. Nevertheless, we observed that the false positive rate remained at zero for the first four tested significance thresholds (5×10⁻⁸, 1.04×10⁻⁷, 1.51×10⁻⁷, and 2.03×10⁻⁷), supporting the robustness of our threshold under the null hypothesis.

We then generated a quantile-quantile (QQ) plot for the p-values of these filtered SNPs. Under the null hypothesis, p-values are expected to follow a uniform distribution^27^, which should result in the points lying along the diagonal (x = y) line in the QQ plot.

### Annotation

Using the new significance threshold of p = 2.03×10^-7^, we uploaded the summary statistics only for the LFVs into the Functional Mapping and Annotation of Genome-Wide Association Studies (FUMA) tool^29^. We used the SNP2GENE module to map genes to the newly significant SNPs. This included positional mapping (capturing all genes within 10 kb of each significant SNP) and eQTL mapping (to link SNPs to genes likely to have their expression affected by that SNP). For all mapped genes, we also checked the corresponding SFARI^28^ and EAGLE^30^ score (if available), which indicates the strength of each gene’s association with autism based on prior studies^28^. However, we note that genes identified using this approach may represent novel biological insights into the genetic architecture of autism and may not yet be captured in curated databases such as SFARI or EAGLE.

### Investigation of the regulatory potential of candidate loci

To further investigate the biological relevance of newly identified ASD-associated low-frequency variants, we queried GTEx^31^ for expression quantitative trait loci (eQTL) associations. Regulatory potential was assessed using the SCREEN (Search Candidate cis-Regulatory Elements by ENCODE)^32,33^ database. For variants lacking direct regulatory annotation, we used the LDproxy tool in the LDlink database^34^ to identify proxy SNPs in strong LD (R² > 0.6) that overlap with regulatory elements, leveraging 1000 Genomes Project^24^ European reference data.

### Gene Expression Analysis

ASD candidate genes prioritized by common and rare variants are often found to be enriched in genes related to synapse, transcription/translation regulation, chromatin organization, and have brain-specific expressions ^11,35,36^. We hypothesized that genes prioritized by LFVs should share the same theme of pathways and expression patterns. We obtained gene expression data at both the tissue level and the cell-type level from the Human Protein Atlas (HPA) database^37^. The tissue-level expression data (bulk RNA-seq) were compiled from HPA *in-house* dataset together with Genotype-Tissue Expression Consortium (GTEx) and normalized into a consensus expression dataset, which contained data for 50 different tissues, capturing 20,162 genes. Similarly, the cell-type-level expression data (single-cell RNA-seq; scRNA-seq) obtained from HPA were compiled from multiple studies including the Single Cell Expression Atlas ^38^, the Human Cell Atlas^39^, the Gene Expression Omnibus^40^, the Tabula Sapiens^41^, the Allen Brain Atlas^42^ and the European Genome-phenome Archive^43^ and combined into a consensus dataset. This consensus dataset contained expression levels of 20,282 genes in 81 cell types, including immune cells, glial cells, muscle cells, and neuronal cells. To be able to compare the expression profile across tissues or cell types, a normalized Transcripts Per Million (nTPM) value within tissue type or cell-type cluster was calculated for each gene using the trimmed mean of M value (TMM) as a normalizing factor. The nTPM value of each gene tagged by associated SNPs using the threshold of p = 2.03×10-7 was compared between each tissue or cell-type to the rest using Wilcoxon’s rank sum test. Given the limited statistical power when using the tissue-level or cell-type-level summarized nTPM values over the sample-level expression profile, we did not perform any multiple testing correction. We are reporting here the nominally significant results with Wilcoxon’s one-sided p<0.05 when comparing if a gene expression level of a particular tissue or cell type is higher than that of the mean expression level of the remaining tissues or cell types.

### Gene Set Enrichment Analysis

We performed gene set enrichment analysis to understand the general function of the new candidate genes identified using the new significance threshold. To improve the statistical power on the gene set enrichment analysis, we extended the list of genes tagged by associated SNPs using GeneMania v3.5.3 by 100 genes^44^. We then performed enrichment analysis with this extended gene list, using g:Profiler^45^. We visualized clusters of enriched gene sets using Cytoscape v3.10 and EnrichmentMap plugin v3.5.0^46^. The cluster labels were annotated using AutoAnnotate plugin v1.5.1.

## Results

### Determining an Appropriate Significance Threshold for Low-Frequency Variants

Based on previously published summary statistics^11^ variants passing the conventional genome-wide significance threshold of 5×10^−8^ were considered significant. Under this threshold, none of the low-frequency variants (LFvar) reached significance, while 93 common variants (MAF > 5%) did (Figure 1). To better assess the detection of LFvar signals, we evaluated a range of alternative significance thresholds derived from Bonferroni correction using the different LD cut-offs for LFVar pruning (see Section: *Significance threshold inference based on Bonferroni correction*) and compared them against the conventional genome-wide and suggestive thresholds ^47^ based on their proportion of non-effect variants detected- and the proportion of true-effect variants missed (Figure 2).

Using our simulation-based approach, we systematically varied the significance threshold to evaluate the balance between detecting non-effect variants and missing true-effect variants. We identified 2.03×10⁻⁷ as the optimal significance threshold, i.e. the most stringent threshold at which sensitivity for true-effect variants improved, while rates of False Positive did not increase. (Figure 2-C, Table S1).

To ensure that this new threshold maintains appropriate control of false positives, we verified this by applying our approach on a set of SNPs selected for their low likelihood of being associated with ASD. As expected under the null hypothesis, the p-values for these SNPs followed a uniform distribution, resulting in a linear Q-Q plot. The absence of p-values deviating above the x = y line (Figure 2-B) indicates no excess detection of non-effect variants and suggests the revised threshold is appropriately calibrated.

### Gene mapping

Using the Bonferroni-corrected significance threshold of 2.03×10^-7^ we detected three LFvar that were not detected with the standard genome-wide significance threshold of 5×10^-8^ (Figure 3A). These include chr7:105103772:A:G,T (rs111931861) (Figure 3B), chr8:48036474:A:G (rs761205017) (Figure 3C), and chr19:36948739:G:A (rs138867053) (Figure 3D; Table S2). Unlike typical common-variant GWAS signals, these LFVs are not consistently surrounded by clusters of near-significant variants. This pattern is expected for LFVs, which tend to exhibit reduced local linkage disequilibrium and are often represented by a limited number of correlated markers^48^. To further investigate their potential biological relevance, we applied both the positional mapping and eQTL mapping modules available on FUMA^29^ (Table S3).

**Figure 3.**
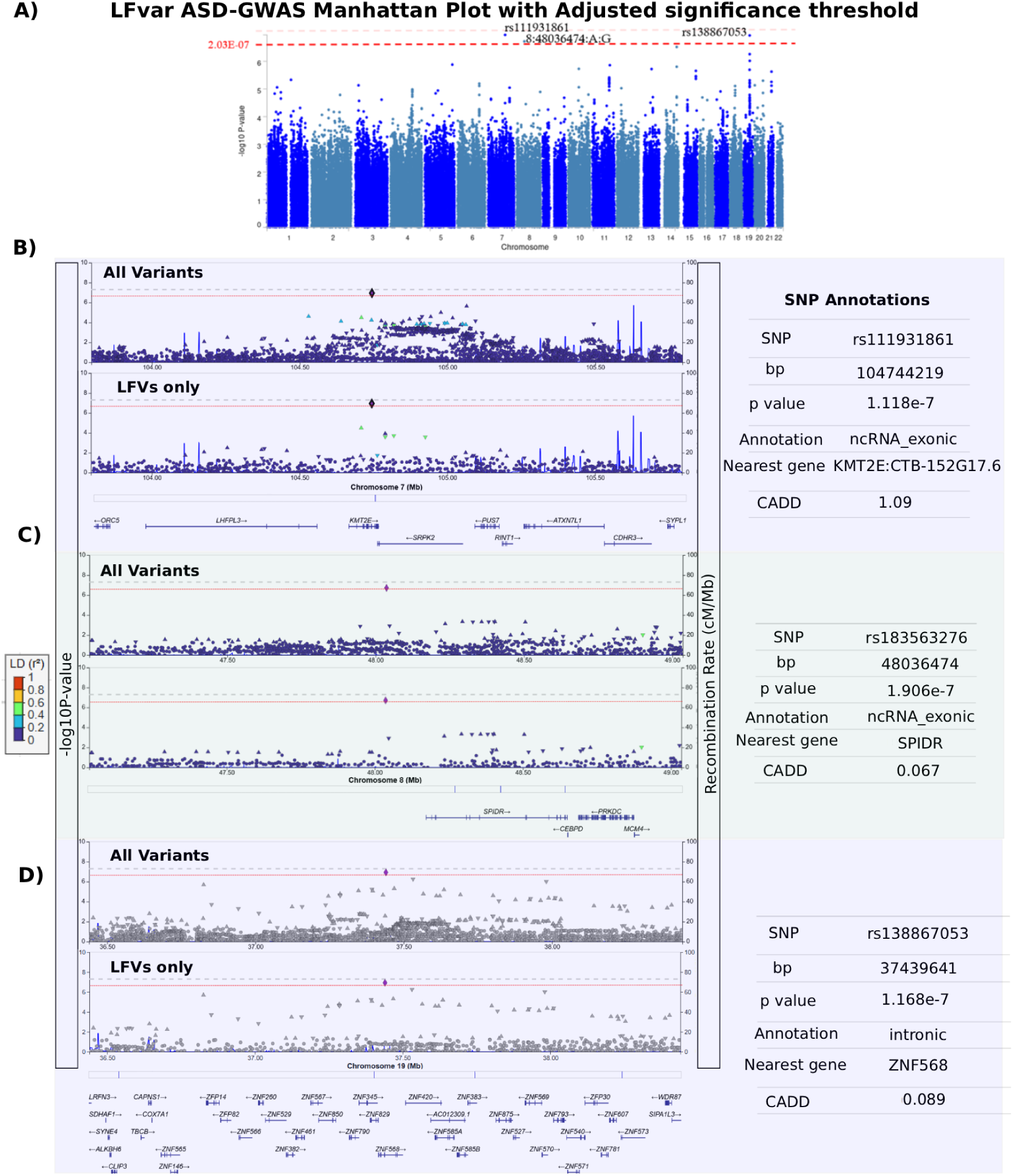
Significant ASD-associated LFVs after adjusting the p value threshold to 2.03×10^-7^. A) Manhattan plot showing the new significance threshold and highlighting variants that surpass this threshold. B) LocusZoom plot of the significant locus on chromosome 7, with annotations for the lead SNP rs111931861 shown on the right. C) LocusZoom plot of the significant locus on chromosome 8, highlighting the lead SNP chr8:48036474:A:G, annotated on the right. D) LocusZoom plot of the significant locus on chromosome 19, with annotations for the lead SNP rs138867053 displayed on the right. In each LocusZoom plot, the lead SNP is shown in purple, and surrounding SNPs are coloured according to their linkage disequilibrium (LD) with the lead SNP, using a gradient where warmer colours indicate stronger LD.

rs111931861, located on chromosome 7, is in proximity of both *KMT2E* and *SRPK2* through positional and eQTL mapping methods. The gene *KMT2E* was previously associated with ASD based on rare variant studies, including copy number loss^49,50^, frameshift variants^50–52^, inversions^53^, intronic variants^54,55^, missense variants^51^, and stop gain variants^51^. Due to these associations, *KMT2E* currently holds a SFARI^28^ score of 1S. The score of 1 indicates a high confidence of ASD association, and the S (syndromic) category indicates that variations in the gene are also linked to additional characteristics not required for an ASD diagnosis. *KMT2E* is also linked to intellectual disability and epilepsy^50^. The second variant, rs761205017, identified on chromosome 8, was positionally mapped to four genes: *SPIDR*, *PRKDC*, *MCM4*, and *UBE2V2*. Of these, *PRKDC* and *MCM4* are listed in the SFARI database^28^, both with a score of 2, indicative of being strong candidates for ASD. The SFARI score for *PRKDC* is supported by associations with 17 previously identified rare variants ^54,56–61^, while the score for *MCM4* is based on four rare variant associations, including one missense variant ^55^, two synonymous variants ^54,55^, and one stop-gain variant^62^. On chromosome 19 (rs138867053), no genes were identified by both gene mapping methods (positional and eQTL) simultaneously. *SIPA1L3*, *SPRED3*, *COX7A1*, *CAPNS1*, *ZNF875*, *ZFP30*, and *ZNF781* were mapped by the eQTL method, while *ZNF420*, *ZNF568*, and *ZNF570* were identified through positional mapping alone.

### Genes expression in relevant ASD tissues and gene set enrichments

GWAS analysis identified potential genes or loci implicated in disease traits by detecting tagging variants with significant associations. Our analysis revealed three additional variants: rs111931861 on chromosome 7, rs761205017 on chromosome 8, and rs138867053 on chromosome 19. All three are located in noncoding regions. GTEx^31^ eQTL data suggests that these variants may be associated with altered transcript levels of nearby genes (Table 1).

**Table 1.** Summary of regulatory annotation and linkage disequilibrium (LD) information for ASD-associated low-frequency variants and their LD proxies. The table lists the key ASD-associated low-frequency variants (“Searching variants”) and their most strongly linked proxy SNPs (“Tagged variants”), as identified through LD analysis. Chromosomal location (Chr), LD measures (R² and D’), and functional annotations from FORGEdb^63^ and RegulomeDB^64^ are provided. LD values were calculated based on European populations from the 1000 Genomes Project. “NA” indicates that no suitable tagging variant was identified within the LD thresholds applied.

| Chr | Searching variants | Tagged variants | R2 | D' | FORGEdb | RegulomeDB |
| --- | --- | --- | --- | --- | --- | --- |
| 7 | rs111931861 | rs74426616 | 0.7851 | 0.945 | 8 | 2b |
|  |  | rs75367503 | 0.6885 | 0.9674 | 7 | 3a |
|  |  | rs77014937 | 0.6885 | 0.9674 | 7 | 3a |
| 8 | rs761205017 | NA | NA | NA | NA | NA |
| 19 | rs138867053 | rs147118670 | 1.0 | 1.0 | 5 | 6 |
|  |  | rs150560882 | 0.7023 | 1.0 | 5 | 5 |
|  |  | rs151298495 | 0.7023 | 1.0 | 8 | 5 |
|  |  | rs141061193 | 0.7023 | 1.0 | 6 | 3a |
|  |  | rs117172205 | 0.6258 | 0.9153 | 4 | 5 |
|  |  | rs79235019 | 0.6258 | 0.9153 | 6 | 7 |

A follow-up query using the SCREEN database indicated that none of these variants reside within annotated regulatory regions. However, further investigation using the LDproxy tool in the LDlink database^34^ identified several regulatory variants in strong linkage disequilibrium (LD) with these three, suggesting potential functional relevance. Thus, by lowering the p-value threshold, these newly identified ASD-associated variants may help explain previously unexplored aspects of disease etiology by uncovering additional genetic mechanisms.

We also investigate the gene expression of the candidate genes tagged by associated SNPs in different tissues (Table S4) and cell types (Table S5). Using gene expression data from Human Protein Atlas^37^, we are reporting here the results with Wilcoxon’s one-sided p<0.05 when comparing if a gene expression level of a particular tissue or cell type is higher than that of the mean expression level of the remaining tissues or cell types. While the majority of the candidate genes do not exhibit brain-specific expression (higher expression in the brain compared to other tissues), *SPIDR*, *SPRED3*, and *UBE2V2* are highly expressed in brain tissues (Figure 4A). Cell-type-level expression showed *SPIDR* and *ZNF420* to be highly expressed in excitatory neurons, while *SRPK2 and UBE2V2* were highly expressed in oligodendrocytes and Müller glia cells, respectively. For other plausible ASD-relevant tissues, we observed 1) *KMT2E* and *MCM4* predominantly expressed in bone marrow and immune cells, ZNF30 and ZNF568 in retina, SRPK2, ZNF420, and ZNF570 in testis, and CAPNS1 and COX7A1 in heart and skeletal muscles (Figure 4A).

**Figure 4.**
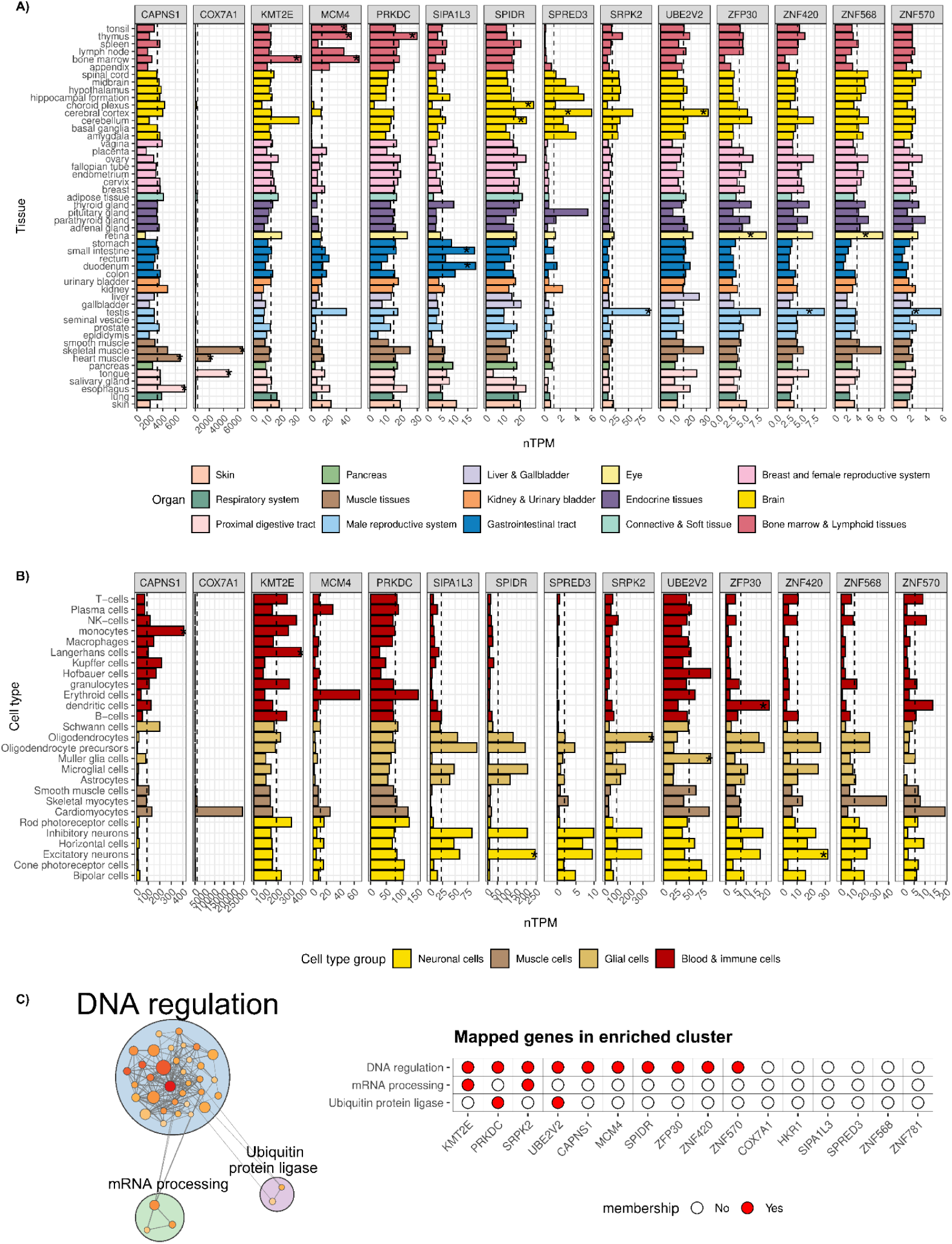
Gene expression level of 14 out of the 16 prioritized genes. A) Tissue-level bulk RNA-seq expression profile. B) Cell-type-level single-cell RNA-seq expression profile. A dashed line indicates mean expression level across tissues or cell-types. An asterisk indicates significantly higher expression level (Wilcoxon’s one-side p < 0.05) compared to the mean expression level. C) Gene set cluster enriched for mapped genes and their 100 extended neighbouring genes. Normalized transcripts per million (nTPM) values allowing across tissues/cell types comparison, were obtained from the Human Protein Atlas database.

In addition to the gene expression analysis of the mapped genes, we also performed g:Profiler gene set enrichment analysis by extending the list of mapped genes by another 100 genes using GeneMania (with equal weight option by data type). The gene set enrichment result was summarized by EnrichmentMap in Cytoscape. We found the gene list to be enriched in basic genome regulation function, i.e., DNA regulation, mRNA processing, and ubiquitin protein ligase (Figure 4 C). We found *KMT2E*, *PRKDC*, *SRPK2*, *UBE2V2*, *CAPNS1*, *MCM4*, *SPIDR, ZFP30*, *ZNF420*, and *ZNF570* to be involved in DNA regulation. Only *KMT2E* and *SRPK2* were in mRNA processing, and only *PRKDC* and *UBE2V2* were in ubiquitin protein ligase (Figure 4C).

Like ASD candidate genes implicated by rare and common variants, we hypothesized that the expression of candidate genes prioritized in this study to be brain-specific, out of the 14 genes with expression profile available, three of them showed brain-specific expression. The gene set enrichment analysis showed that these three genes along with 7 other genes are related to basic DNA/RNA regulation.

## Discussion

In this study, we argue that genome-wide significance thresholds used for GWAS are appropriate for the detection of common SNPs but prevent the identification of association signals driven by variants in the low frequency range. Here, we calculated a genome-wide significance threshold appropriate for LFVs while avoiding inflation of type-1 error. Our findings emphasize that standard genome-wide significance thresholds may cause important genetic contributions from LFVs to be overlooked. Consistent with the original understanding of GWAS thresholds, these statistical benchmarks, while effective in minimizing type 1 errors for common variants, are less suited for detecting associations at lower allele frequencies.

The simulation studies we conducted further support this adjustment, demonstrating a more balanced approach to controlling both false positive and false negative errors. Notably, we observed that the unmodified significance threshold, optimized for common variants, resulted in disproportionately high false negative rates for LFVs. Our MAF-appropriate adjustments successfully reduced this risk while preserving the robustness of the statistical models. These findings are particularly relevant as the field increasingly acknowledges that rare and low-frequency variants are essential components of the genetic architecture underlying complex traits and disorders, such as autism.

Nevertheless, even with MAF-appropriate significance thresholds, the type II error rate for low-frequency variants remains substantial (Figure 2A). This reflects a fundamental limitation of association testing for LFVs, where reduced allele counts inherently limit statistical power, particularly in realistic GWAS sample sizes. While our adjusted threshold substantially improves sensitivity relative to the standard genome-wide significance threshold, it does not fully overcome the power constraints imposed by low allele frequencies.

It is important to acknowledge that our approach does not suggest the abandonment of strict statistical thresholds but rather advocates for nuanced adjustments that reflect the biological and statistical realities of LFVs. The use of adaptive significance thresholds is a natural progression in the evolution of GWAS, one that is critical for capturing the full spectrum of genetic variation influencing neurodevelopmental disorders.

By implementing MAF-appropriate significance thresholds, we identified three novel genomic loci associated with autism, which were linked to genes implicated in basic genome regulation such as histone modification and zinc finger protein (ZNF) clusters, underscoring the possibility that regulatory mechanisms may play a critical role in neurodevelopmental disorders. It is noteworthy that, in contrast to typical common-variant GWAS signals, the associated LFVs identified here are not consistently surrounded by dense clusters of near-significant variants in regional association plots. Such extended LD-based association patterns are a hallmark of common variants that reside in strong linkage disequilibrium LD blocks and are therefore tagged by many correlated SNPs. In contrast, LFVs tend to exhibit reduced local LD and are often represented by a limited number of correlated markers, resulting in more isolated association peaks ^48^.

Through the expression analysis, we highlighted that three genes (*SPIDR*, *SPRED3*, *UBE2V2*) from the two candidate loci (chromosome 8 and chromosome 19 loci) have higher expression levels in the brain compared to the other tissues. Like ASD candidate genes implicated in rare and common variants ^11,65^, this suggests a potential role of LFVs to also be related to the nervous system development. Two genes (*SPIDR* and *ZNF420*) from the same loci also exhibit higher expression in excitatory neurons compared to other brain cell types. Excitatory neurons are the major cell type responsible for synaptic transmission and a single-cell study has shown that excitatory neurons were preferentially affected in ASD individuals compared to controls ^66^. Another gene of interest is *KMT2E*, a well-established ASD candidate listed in the SFARI database^28^. Although *KMT2E* shows preferentially higher expression in immune-related tissues and cell types, its association with ASD may reflect the gene’s pleiotropic functions^67^.

The loci we identified on chromosome 19 are enriched with genes known for their roles in chromatin remodeling and transcriptional regulation. For example, histone modification genes regulate the accessibility of the genome, influencing which genes are expressed in a cell at a given time^68^. Dysregulation of these processes has previously been implicated in neurodevelopmental disorders, including autism, providing a plausible link between genetic variation and altered brain development^69^. However, our results suggest that the influence of regulatory elements may extend beyond histone modification. The large ZNF cluster on chromosome 19 (19q13.12) identified in this study, for instance, is particularly intriguing because ZNFs are key regulators of gene expression, particularly in processes involving developmental timing and cell differentiation.

We hypothesize that LFVs, by their nature, may disproportionately affect these genome regulatory genes and gene networks, potentially disrupting the complex processes that govern human brain development. These variants may have larger effect sizes due to their involvement in fundamental pathways that control the spatial and temporal dynamics of the genome. Given the critical periods of brain development and the intricate choreography of gene expression during these phases, it is plausible that even modest disruptions in regulatory genes could lead to neurodevelopmental abnormalities, including autism^70^. Our results therefore suggest that these basic regulatory mechanisms are not only relevant but may be a key aspect of the genetic architecture underlying autism risk. While some LFVs may be subject to purifying selection, their persistence can also reflect demographic history, polygenic inheritance, or elevated mutation rates, underscoring multiple mechanisms beyond selection alone.

The potential impact of LFVs aligns with evidence that autism-associated variants often lie in non-coding regions. A notable example is the cluster of ZNFs at the 19q13.12 locus, transcription factors involved in gene regulation. Although the role of ZNFs in autism is not well understood, this study suggests they deserve further investigation. Given their roles in cellular differentiation and development, ZNF clusters may offer valuable insights into autism’s genetic basis.

## Conclusion

Our findings highlight the need to reconsider significance thresholds for genome-wide association studies, particularly in the context of LFVs. By calculating a threshold specific for the variants in the low allele frequency (1-5%) range, we demonstrate that previously undetected loci can be identified without increasing the detection of non-effect variants, offering novel insights into the genetic architecture of autism. This work highlights the importance of refining statistical frameworks to ensure that meaningful biological discoveries are not missed due to overly stringent significance criteria. As GWASs continue to expand in scope and complexity, embracing these methodological advancements will be essential for fully elucidating the genomic underpinnings of neurodevelopmental disorders and other complex traits.

In addition, our findings indicate that LFVs may be especially relevant, influencing complex processes like brain development through their impact on gene networks. Future research should focus on further dissecting the role of genome regulatory elements, particularly ZNFs, in autism and related neurodevelopmental disorders, as these genes and their regulatory motifs may hold the key to unlocking the full genetic complexity of these conditions.

## Data Availability

All data produced in the present work are contained in the manuscript

https://pgc.unc.edu/for-researchers/download-results/

## References

1. Grotzinger, A. D. et al. The Landscape of Shared and Divergent Genetic Influences across 14 Psychiatric Disorders. medRxiv: the preprint server for health sciences (2025) doi:10.1101/2025.01.14.25320574.

2. Demontis, D. et al. Genome-wide analyses of ADHD identify 27 risk loci, refine the genetic architecture and implicate several cognitive domains. Nature genetics 55, (2023).

3. Wightman, D. P. et al. A genome-wide association study with 1,126,563 individuals identifies new risk loci for Alzheimer’s disease. Nature genetics 53, (2021).

4. Forstner, A. J. et al. Genome-wide association study of panic disorder reveals genetic overlap with neuroticism and depression. Molecular psychiatry 26, (2021).

5. Watson, H. J. et al. Genome-wide association study identifies eight risk loci and implicates metabo-psychiatric origins for anorexia nervosa. Nature genetics 51, (2019).

6. Genomic Relationships, Novel Loci, and Pleiotropic Mechanisms across Eight Psychiatric Disorders. Cell 179, (2019).

7. Antaki, D. et al. A phenotypic spectrum of autism is attributable to the combined effects of rare variants, polygenic risk and sex. Nature Genetics 54, 1284–1292 (2022).

8. Davies, R. W. et al. Using common genetic variation to examine phenotypic expression and risk prediction in 22q11.2 deletion syndrome. Nature medicine 26, (2020).

9. Bergen, S. E. et al. Joint Contributions of Rare Copy Number Variants and Common SNPs to Risk for Schizophrenia. The American journal of psychiatry 176, (2019).

10. Trost, B. et al. Genomic architecture of autism from comprehensive whole-genome sequence annotation. Cell 185, 4409–4427.e18 (2022).

11. Grove, J. et al. Identification of common genetic risk variants for autism spectrum disorder. Nature Genetics 51, 431–444 (2019).

12. Marshall, C. R. et al. Contribution of copy number variants to schizophrenia from a genome-wide study of 41,321 subjects. Nature Genetics 49, 27–35 (2016).

13. Maihofer, A. X. et al. Rare copy number variation in posttraumatic stress disorder. Molecular Psychiatry 27, 5062–5069 (2022).

14. Fournier, T. et al. Extensive impact of low-frequency variants on the phenotypic landscape at population-scale. (2019) doi:10.7554/eLife.49258.

15. Tick, B., Bolton, P., Happé, F., Rutter, M. & Rijsdijk, F. Heritability of autism spectrum disorders: a meta-analysis of twin studies. J Child Psychol Psychiatry 57, 585–595 (2016).

16. Klei, L. et al. How rare and common risk variation jointly affect liability for autism spectrum disorder. Mol Autism 12, 66 (2021).

17. Lin, D. Y. & Zeng, D. On the relative efficiency of using summary statistics versus individual-level data in meta-analysis. Biometrika 97, 321–332 (2010).

18. Xing, G., Lin, C.-Y., Wooding, S. P. & Xing, C. Blindly using Wald’s test can miss rare disease-causal variants in case-control association studies. Ann Hum Genet 76, 168–177 (2012).

19. Fadista, J., Manning, A. K., Florez, J. C. & Groop, L. The (in)famous GWAS P-value threshold revisited and updated for low-frequency variants. European Journal of Human Genetics 24, 1202–1205 (2016).

20. Download Results – PGC. https://pgc.unc.edu/for-researchers/download-results/.

21. Purcell, S. et al. PLINK: a tool set for whole-genome association and population-based linkage analyses. Am J Hum Genet 81, 559–575 (2007).

22. Enrichment analyses identify shared associations for 25 quantitative traits in over 600,000 individuals from seven diverse ancestries. The American Journal of Human Genetics 109, 871–884 (2022).

23. Mendes, M. et al. Chromosome X-wide common variant association study in autism spectrum disorder. Am J Hum Genet 112, 135–153 (2025).

24. A global reference for human genetic variation. Nature 526, 68–74 (2015).

25. Gauderman, W. J. et al. Pathway polygenic risk scores (pPRS) for the analysis of gene-environment interaction. PLoS Genet 21, e1011543 (2025).

26. Puig, X., Ginebra, J. & Graffelman, J. Bayesian model selection for the study of Hardy-Weinberg proportions and homogeneity of gender allele frequencies. Heredity (Edinb) 123, 549–564 (2019).

27. Wang, J., Yu, J., Lipka, A. E. & Zhang, Z. Interpretation of Manhattan Plots and Other Outputs of Genome-Wide Association Studies. Genome-Wide Association Studies 63–80 (2022).

28. Abrahams, B. S. et al. SFARI Gene 2.0: a community-driven knowledgebase for the autism spectrum disorders (ASDs). Mol Autism 4, 36 (2013).

29. Watanabe, K., Taskesen, E. & van Bochoven, A. Functional mapping and annotation of genetic associations with FUMA. Nature Communications 8, 1–11 (2017).

30. Schaaf, C. P. et al. A framework for an evidence-based gene list relevant to autism spectrum disorder. Nat Rev Genet 21, 367–376 (2020).

31. Lonsdale, J. et al. The Genotype-Tissue Expression (GTEx) project. Nature Genetics 45, 580–585 (2013).

32. ENCODE Project Consortium et al. Expanded encyclopaedias of DNA elements in the human and mouse genomes. Nature 583, 699–710 (2020).

33. SCREEN: Search Candidate cis-Regulatory Elements by ENCODE. https://screen.wenglab.org/about.

34. Machiela, M. J. & Chanock, S. J. LDlink: a web-based application for exploring population-specific haplotype structure and linking correlated alleles of possible functional variants. Bioinformatics 31, 3555–3557 (2015).

35. De Rubeis, S. et al. Synaptic, transcriptional and chromatin genes disrupted in autism. Nature 515, 209–215 (2014).

36. Pinto, D. et al. Functional impact of global rare copy number variation in autism spectrum disorders. Nature 466, 368–372 (2010).

37. Karlsson, M. et al. A single-cell type transcriptomics map of human tissues. Science advances 7, (2021).

38. Single Cell Expression Atlas. https://www.ebi.ac.uk/gxa/sc/home.

39. Home page. https://www.humancellatlas.org/.

40. Home - GEO - NCBI. https://www.ncbi.nlm.nih.gov/geo/.

41. Tabula Sapiens. Tabula Sapiens https://tabula-sapiens.sf.czbiohub.org/.

42. Brain Map - brain-map.org. https://portal.brain-map.org/.

43. EGA European Genome-Phenome Archive - EGA European Genome-Phenome Archive. https://www.ebi.ac.uk/ega/.

44. Warde-Farley, D. et al. The GeneMANIA prediction server: biological network integration for gene prioritization and predicting gene function. Nucleic acids research 38, (2010).

45. Reimand, J. et al. g:Profiler-a web server for functional interpretation of gene lists (2016 update). Nucleic acids research 44, (2016).

46. Merico, D., Isserlin, R., Stueker, O., Emili, A. & Bader, G. D. Enrichment map: a network-based method for gene-set enrichment visualization and interpretation. PloS one 5, (2010).

47. Duggal, P., Gillanders, E. M., Holmes, T. N. & Bailey-Wilson, J. E. Establishing an adjusted p-value threshold to control the family-wide type 1 error in genome wide association studies. BMC Genomics 9, 1–8 (2008).

48. Wang, D. et al. cLD: Rare-variant linkage disequilibrium between genomic regions identifies novel genomic interactions. PLoS Genet 19, e1011074 (2023).

49. Velmans, C. et al. O’Donnell-Luria-Rodan syndrome: description of a second multinational cohort and refinement of the phenotypic spectrum. J Med Genet 59, 697–705 (2022).

50. O’Donnell-Luria, A. H. et al. Heterozygous Variants in KMT2E Cause a Spectrum of Neurodevelopmental Disorders and Epilepsy. Am J Hum Genet 104, 1210–1222 (2019).

51. Wang, T. et al. Large-scale targeted sequencing identifies risk genes for neurodevelopmental disorders. Nat Commun 11, 4932 (2020).

52. Abreu, N. J. et al. Novel truncating variant in associated with cerebellar hypoplasia and velopharyngeal dysfunction. Clin Case Rep 10, e05277 (2022).

53. Pagnamenta, A. T. et al. The impact of inversions across 33,924 families with rare disease from a national genome sequencing project. Am J Hum Genet 111, 1140–1164 (2024).

54. Zhou, X. et al. Integrating de novo and inherited variants in 42,607 autism cases identifies mutations in new moderate-risk genes. Nat Genet 54, 1305–1319 (2022).

55. Fromer, M. et al. De novo mutations in schizophrenia implicate synaptic networks. Nature 506, 179–184 (2014).

56. Lim, E. T. et al. Rates, distribution and implications of postzygotic mosaic mutations in autism spectrum disorder. Nat Neurosci 20, 1217–1224 (2017).

57. Woodbine, L. et al. PRKDC mutations in a SCID patient with profound neurological abnormalities. J Clin Invest 123, 2969–2980 (2013).

58. Iossifov, I. et al. The contribution of de novo coding mutations to autism spectrum disorder. Nature 515, 216–221 (2014).

59. Callaghan, D. B. et al. Whole genome sequencing and variant discovery in the ASPIRE autism spectrum disorder cohort. Clin Genet 96, 199–206 (2019).

60. Feliciano, P. et al. Exome sequencing of 457 autism families recruited online provides evidence for autism risk genes. NPJ Genom Med 4, 19 (2019).

61. Woodbury-Smith, M. et al. Mutational Landscape of Autism Spectrum Disorder Brain Tissue. Genes (Basel) 13, (2022).

62. Cirnigliaro, M. et al. The contributions of rare inherited and polygenic risk to ASD in multiplex families. Proc Natl Acad Sci U S A 120, e2215632120 (2023).

63. Breeze, C. E. et al. FORGEdb: a tool for identifying candidate functional variants and uncovering target genes and mechanisms for complex diseases. Genome Biology 25, 1–13 (2024).

64. Boyle, A. P. et al. Annotation of functional variation in personal genomes using RegulomeDB. Genome Res 22, 1790–1797 (2012).

65. Pinto, D. et al. Convergence of genes and cellular pathways dysregulated in autism spectrum disorders. Am J Hum Genet 94, 677–694 (2014).

66. Velmeshev, D. et al. Single-cell genomics identifies cell type-specific molecular changes in autism. Science 364, 685–689 (2019).

67. Heuser, M. et al. Loss of MLL5 results in pleiotropic hematopoietic defects, reduced neutrophil immune function, and extreme sensitivity to DNA demethylation. Blood 113, 1432–1443 (2009).

68. Gibney, E. R. & Nolan, C. M. Epigenetics and gene expression. Heredity 105, 4–13 (2010).

69. Rangasamy, S., D’Mello, S. R. & Narayanan, V. Epigenetics, Autism Spectrum, and Neurodevelopmental Disorders. Neurotherapeutics 10, 742 (2013).

70. Trevino, A. E. et al. Chromatin and gene-regulatory dynamics of the developing human cerebral cortex at single-cell resolution. Cell 184, 5053–5069.e23 (2021).

