## Supplementary Information for "Genome-Wide Significance Reconsidered: Low-Frequency Variants and Regulatory Networks in Autism"

**Affiliations**

**Table S1.** Summary statistics from the simulated genome-wide association study (GWAS), including effect estimates (Outcome), standard errors (std), P-values, log-transformed P-values (logp), absolute effect sizes (absbeta), and prediction metrics (pred, no_effect, range).

**Table S2.** Genomic risk loci identified through FUMA analysis. Columns include the genomic locus identifier, unique variant ID, rsID, chromosome number (chr), base-pair position (pos), association p-value, genomic start and end positions defining the locus, and counts of total, GWAS-significant, independent, and lead SNPs within each locus.

**Table S3.** Gene-based annotation results from FUMA analysis. Genes were mapped to GWAS-derived genomic loci based on positional overlap, eQTL colocalization.

**Table S4.** Gene expression levels across human tissues for FUMA-annotated genes. Expression (nTPM) values for genes of interest were extracted from Protein Atlas and grouped by tissue and organ type.

**Table S5.** Cell type–specific gene expression profiles for FUMA-annotated genes. Expression values (nTPM) are shown for each gene across brain cell types, grouped by major cell type classes.
